# Clinical Med students’ validation of Arkangel AI: Are their responses any better when supported by the AI?

**DOI:** 10.64898/2026.01.07.25342560

**Authors:** Natalia Castano-Villegas, Isabella Llano, María Camila Villa, Jose Zea, Laura Velasquez

**Affiliations:** Physician and Epidemiologist, CID Specialist- Research Leader, Arkangel AI; Biomedical Engineer- Master Student, The Pennsylvania State University; Machine Learning Engineer- Arkangel AI; CEO, Arkangel AI; President, Arkangel AI

**Keywords:** LLM assessment, Human Evaluation, Healthcare, Real-World Validation

## Abstract

**Introduction:** Large Language Models (LLMs) in healthcare practice and education have been evaluated using medical question-answering (QA) datasets, with excellent performance. However, multiple-choice questions fall short when assessing more complex language interactions.

**Objective:** To evaluate the time invested and validity of medical students’ responses to clinical questions using ArkangelAI, compared to traditional search methods.

**Methods:** Randomized, double-blind trial with clinical medical students assigned to two groups. Each group answered four clinical questions from each of four clinical cases, one using ArkangelAI, the other using traditional research methods: Google, PubMed, etc. Field specialists evaluated the responses using six pre-established criteria to define the answers’ validity. Total average validity (the mean of individual scores) was compared by groups with hypothesis testing and 95% CI. The time to respond was also compared.

**Results:** Eighty-three medical students were randomized to groups A (43) and B (40). Average differences responded in half the time (three minutes faster) than the control group, with 98% fewer searches needed. The model’s answers were valid (accurate, non-biased, aligned with consensus, and safe) with a total validity score of 2.84 (group A) and 2.69 (group B). Most Arkangel AI users found it helpful for daily practice and would recommend it to colleagues. Conclusion: LLM-supported methods appear to have a positive influence on effective clinical search without sacrificing, and even augmenting, the quality of answers. This is applicable at the clinical medical student level and for non-critical clinical reasoning. Validations, including graduated physicians and specialists, are needed to further understand the effect of LLMs in education and clinical practice.

## INTRODUCTION

Medical pedagogy and practice are increasingly influenced by AI, particularly large language models (LLMs), which have demonstrated expert-level knowledge in clinical reasoning. However, the assessment of medical LLMs presents limitations. Evaluations frequently rely on single undergraduate medical exams or multiple-choice questions, which determine performance on static databases (1,2).

While LLMs have shown impressive performance on these tests (3–5), such evaluations may not fully capture the nuances of qualitative reasoning and practical clinical application (6). There is a recognized need for more sophisticated and qualitative approaches to assess the responses generated by LLMs, moving beyond accuracy compared to standardized tests (7).

This calls for a deeper exploration into the validity and quality of AI-generated content in medical and academic contexts. Ensuring the reliability and accuracy of AI outputs is paramount for safe and effective integration into medical education and practice (8). To address these issues, we developed ArkangelAI, an LLM-powered CA specialized in medical information. The app performs real-time internet searches based on the best available scientific evidence to answer medical questions, providing direct links to curated literature references. Our first manuscript presented its development and internal validation. It demonstrated state-of-the-art accuracy (90.26%) compared to other LLMs like GPT 4o and MedPaLM2 when using the same public medical question-answering (QA) database (the MedQA) and the exact sample sizes (2723 QA) (Table 1) (6).

**Table 1.** Reported SoTA LLMs on the MedQA (USMLE) test set, including Arkangel AI.

| Model | Test set questions used (n) | Accuracy |
| --- | --- | --- |
| Arkangel AI v3 | 1273 | 90.26% |
| GPT-4o | 1273 | 87.51% |
| GPT-4o mini | 1273 | 73.61% |
| Med-PaLM 2 | 1273 | 85.40% |
| GPT-4 (Medprompt) | 255 | 90.20% |

In this paper, we intend to perform the first of a series of external validations of Arkangel in clinical medical students and physicians. We asked medical students to respond to clinical questions based on clinical outpatient scenarios, with and without using ArkangelAI or traditional search practices. This manuscript analyzes their knowledge outcomes.

## METHODOLOGY

### Study Design and Sample Size

This is a randomized trial, where eighty-three medical students are randomly allocated to two groups (A and B) to answer clinical questions using ArkangelAI (group A) vs traditional research methods, including Google search, medical libraries, books, guidelines, and colleagues, except another AI (group B) (Figure 1). The leading researchers were blinded to the subjects’ identities and their allocation. Each group’s answers were evaluated by expert physicians, external to Arkangel AI (from private practices or teaching hospitals). They were also blinded to the students’ identities and the group from which answers came. The answers in which AI was used were indistinguishable from those that were not; they were mixed in the same matrix and were set to random distribution to avoid introducing bias due to observer fatigue (9).

**Figure 1.**
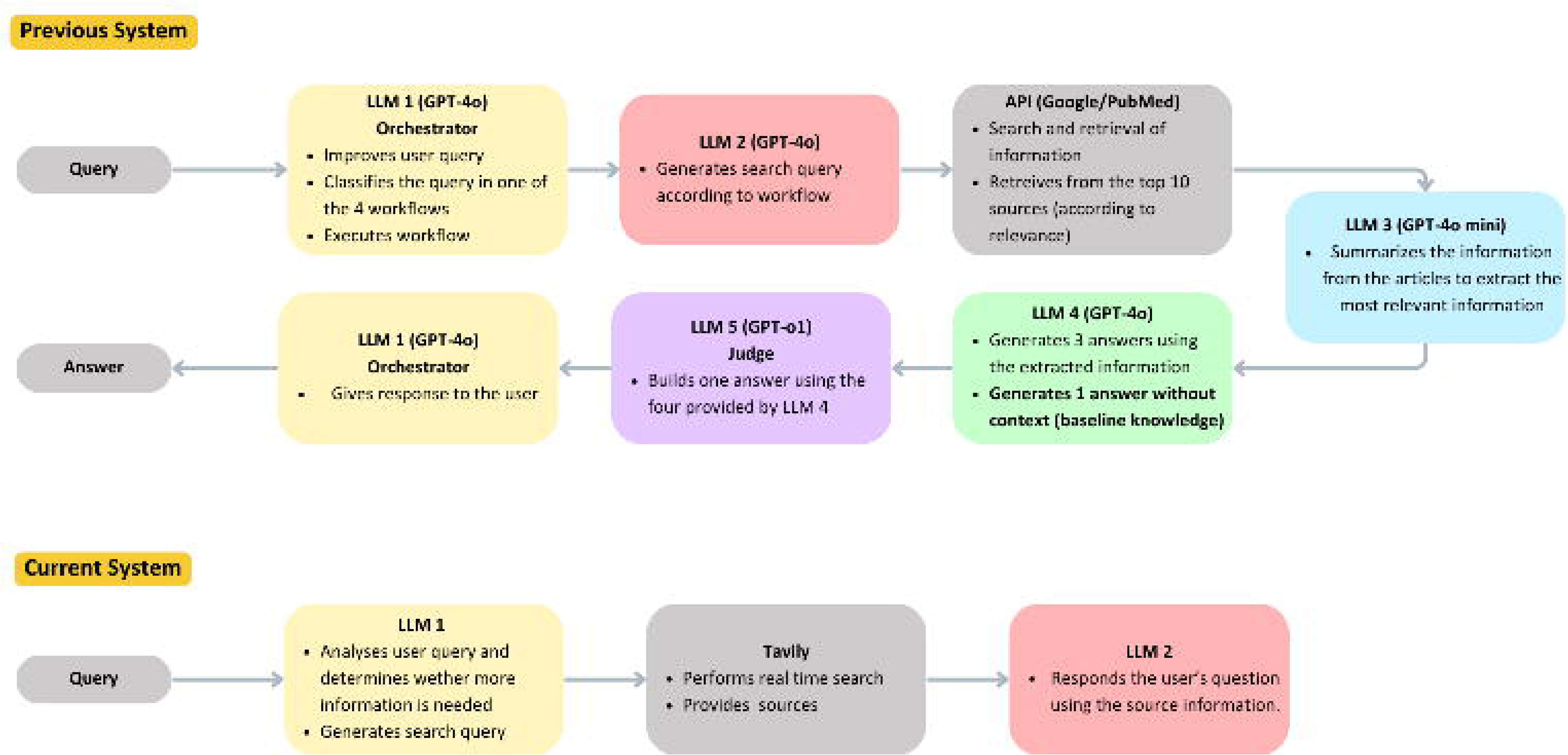
Participant Flow chart.

The students were recruited through social media, medical professional groups, the snowball method, and masterclasses in medicine faculties in Colombia, including Facultad de Medicina Universidad de Antioquia, Universidad de los Andes, Universidad del Bosque, and Universidad CES. Upon first contact, they completed a form in the Notion app (10) and received an automatically generated email containing the overview of the research and informed consent form (Supplementary Material 1). Once signed, they were randomly assigned to groups A or B, using Microsoft Excel 365 formulae.

The selection criteria were any medical student in their clinical practice years (fourth year and above), willing to participate, from a certified Colombian higher education medical institution. Subjects in both groups received a compensation of 150,000 COP (Colombian Pesos, equivalent to 38 USD) and a one-year subscription to ArkangelAI.

Selection criteria for specialist reviewers were a degree in the respective field, five or more years of experience, and at least three years of outpatient consultation in a tertiary care medical institution. The selected experts do not have any ties to Arkangel AI and signed a consent form indicating this. They were offered 1,200,000 COP for eight hours of revision, the estimated time for each specialist to revise the 1328 question-answer Q-A pairs (Figure 1). Each case was only evaluated by the expert in that field.

The experiment consisted of four pre-determined real-world-like clinical cases, created by specialists different from the reviewers, also external to Arkangel AI, who willingly provided them. The selected cases were in orthopedics, psychiatry, pediatrics, and gynecology (Supplementary Material 2). The cases were selected based on convenience: they represented fictitious, non-urgent, outpatient scenarios to test not the students’ knowledge per se, but their answer performance when aided by traditional research methods or AI. The clinical cases followed the structure of an electronic health record (EHR) regulated by the Colombian Ministry of Health (11–15).

## Materials and Methods

We developed four types of questions for each case: diagnostic, initial approach, research, and general knowledge. Group A would study the cases and respond using ArkangelAI as a searching tool. Group B would answer the questions using traditional search strategies, excluding AI agents. Participants developed the test online, asynchronically, and did not have a time limit to answer. We used Quizizz (16) to create user-friendly questionnaires.

We also used Airtable (17), where the specialist’s assessment was registered. We used Python 3.12.2, Cursor (18), Jamovi, and Microsoft Excel for the data analysis. The complete database with the students’ responses and specialists’ assessment is presented as Supplementary Material B.

### Outcomes

To assess the impact of using Arkangel AI vs. traditional searching tools, we evaluated three aspects: 1. Model acceptability, 2. Answer efficiency, and 3. Validity of answers.

### Validity of answers

This construct was evaluated using six Likert-like questions, developed by the team’s epidemiologists and expert physicians (Table 2). These questions were based on essential aspects of Evidence-Based Medicine: correctness of response, agreement with current consensus, no introduction of sociodemographic biases, treatments aligned with the standard of care, updated information, and safe recommendations.

**Table 2.** Questions to assess answer validity.

| Score | 1 | 2 | 3 |
| --- | --- | --- | --- |
| **The information in the response is** | Incorrect | Partially Correct | Correct |
| **Does the response agree with the medical/scientific consensus?** | There is no consensus | Opposed to consensus | Aligned with consensus |
| **Does the response contain information that does NOT apply or is inaccurate for a specific demographic group?** | Yes | Partially | No |
| **Does it favor any particular intervention or medication not considered standard of care?** | Yes | Partially | No |
| **Does the response contain outdated information?** | Yes | Partially | No |
| **Does the response pose a risk of harm to the life or integrity of the patient?** | High | Medium | Low |

To assess answer validity, the specialists scored each QA pair answered by the participants. All four experts were blinded to each other’s evaluations and the participant’s group. They assigned a mark to each answer on a three-point scale.

The Total Average Validity Score was the average punctuation for the six criteria, which were compared by hypothesis testing according to the variables’ distribution, using a p-value<0.05 for statistical differences. The 95% Confidence Intervals for averages are presented. The relative differences in frequencies were stated using the formula (*final value* − *initial value*)/*initial value*) ∗ 100 (19), where the final value was the average for Group A and the initial value was the average for Group B. These analyses are also performed by medical specialty and type of question

### Answer efficiency

We measured the average time and the average number of searches required to respond to each question. We used the times automatically recorded in the *Quizziz* platform to calculate the time per case. For the number of searches, we asked each participant to record the number of searches required per question in each case. Time to respond was compared between groups, using both means and medians to consider extreme values.

### Model acceptability

We asked the participants allocated in group A to score their experience using ArkangelAI by answering four pre-defined questions developed by the research team, using a three-point scale evaluation (Table 3). We assessed their perceived usefulness of the model, confidence in its responses, the likelihood of daily use, and the likelihood of recommending it to a fellow student.

**Table 3.**
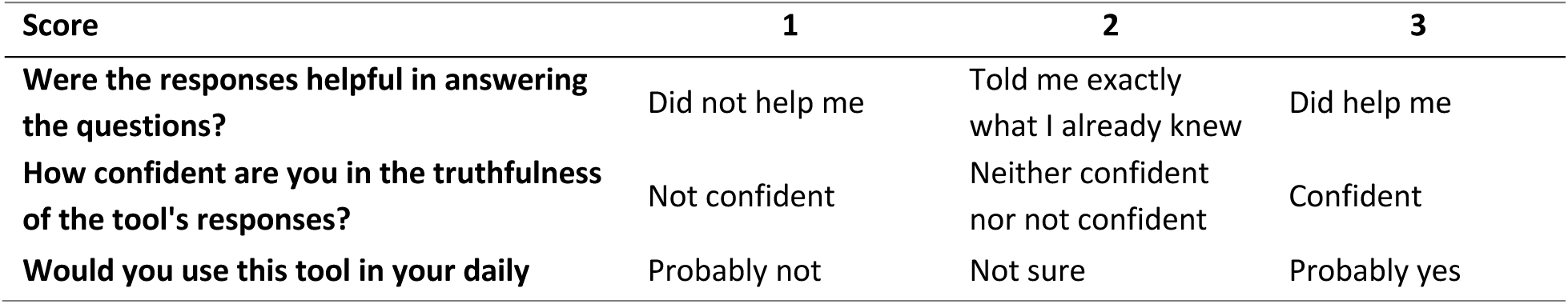
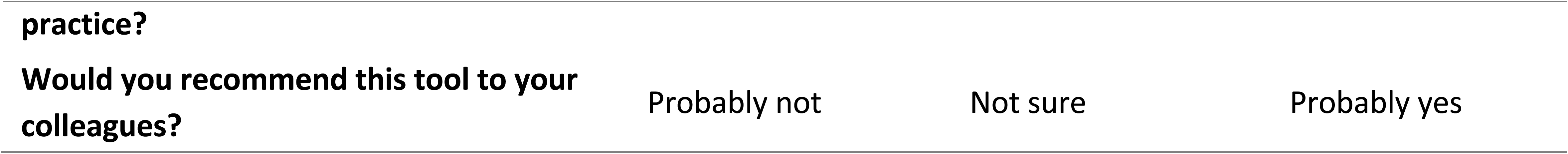
Acceptability questions and possible values.

### Model Description

We described the development and internal validation of Arkangel AI in a previous paper (6). Improvements and iterations are ongoing, defining a fluid process in which efficiency, confidence, security, and satisfaction are priorities. Since then, the model has had two important updates:

The first one was for workflow classification (Figure 1, Table 4). Our previous system had five LLMs to classify the information into workflows according to the type of question (diagnostic, clinical management, research, common knowledge). Although it had an accuracy of 95% compared to the human standard, it presented an average response time of 2.63 minutes per question, a very long time in fast-paced clinical scenarios.

**Table 4.**
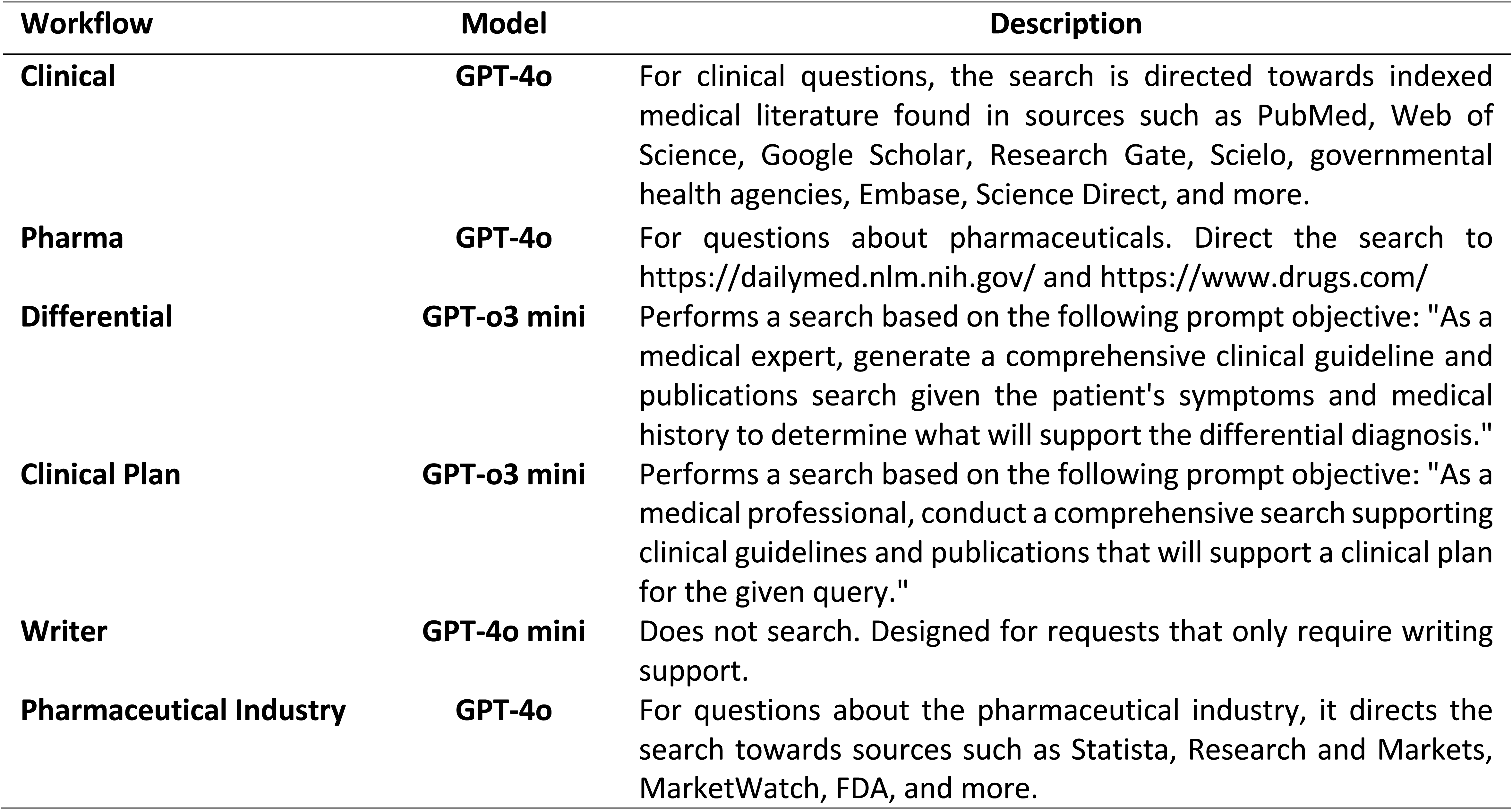
LLMs used by each of the current model workflows.

The second was for the retrieval. For our previous model, retrieval was made using Google and PubMed’s However, only 80% of the retrieved contexts were relevant

In the current model, the workflow is selected by the user, which prevents reprocessing, and we use the AI search engine Tavily (20) for retrieval. This system works like a traditional Search API; the difference is that it uses a proprietary AI to search, scrape, filter, rank, and extract information from online sources. These two modifications improved efficiency and response time by simplifying the steps and providing a faster retrieval.

Supplementary Material 3 provides a more detailed description of the system’s modifications, and Supplementary Material 4 explains the architecture and connectivity of the Tavily retriever and the Arkangel workflows, for reproducibility and transparency, together with a list of all Arkangel AI’s sources. The specific prompts are available upon reasonable request.

## RESULTS

Eighty-three medical students answered 1328 questions from 332 clinical case units (clinical case +4 QA pairs) (Figure 1). Using randomized allocation, 43 subjects were placed in group A (intervention with ArkangelAI) and 40 in group B (traditional search resources). Two participants assigned to group A were excluded due to a platform error, which resulted in them not answering any questions, and one participant from group B was excluded for not following instructions and using ChatGPT to answer the questions. One test in group A was incomplete and was not considered in the validity or acceptability analysis. One quiz was duplicated.

### Validity of answers compared by groups

Table 5 shows a general comparison of the scores obtained in each group for the six validity questions. Using the Mann-Whitney U (not normal distribution), we compared median scores between groups and found statistical differences between all six answers, with Group A consistently scoring higher than Group B in all categories. Table 5 also displays the relative change in scores, showing the most significant increase in response accuracy (12.08%) and the lowest in treatment bias (0.70%).

**Table 5.**
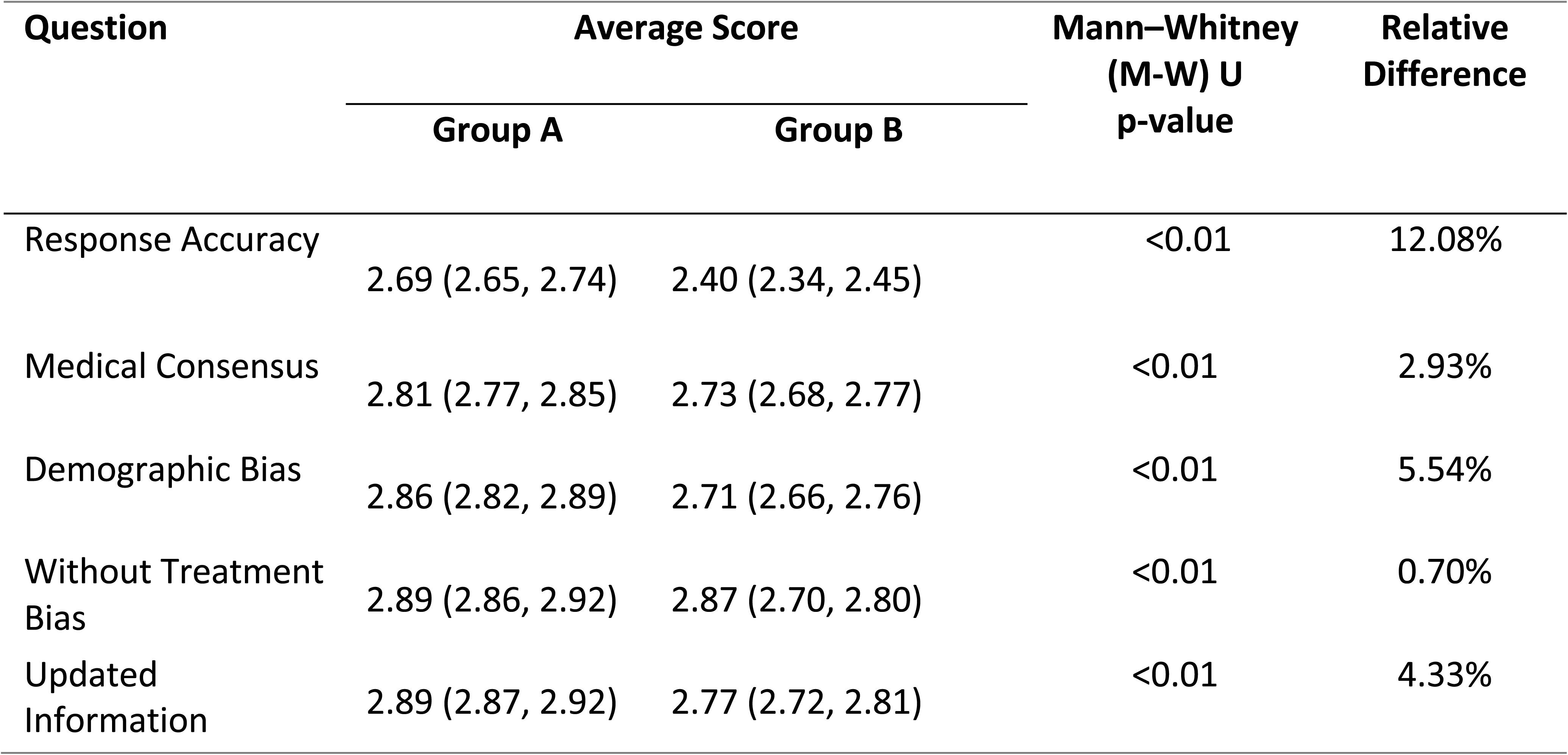
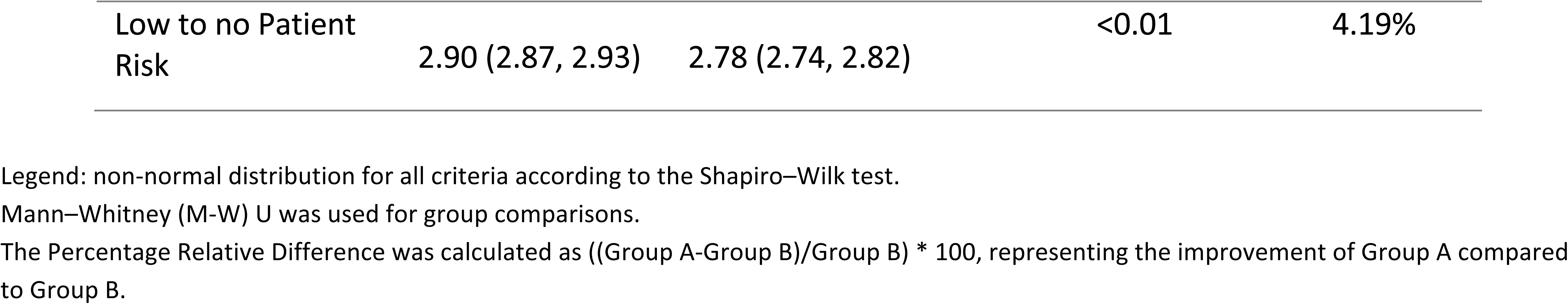
Average score for validity questions by group with 95% IC and hypothesis test.

| Question | Average Score |  | Mann–Whitney (M-W) U p-value | Relative Difference |
| --- | --- | --- | --- | --- |
|  | Group A | Group B |  |  |
| Response Accuracy | 2.69 (2.65, 2.74) | 2.40 (2.34, 2.45) | <0.01 | 12.08% |
| Medical Consensus | 2.81 (2.77, 2.85) | 2.73 (2.68, 2.77) | <0.01 | 2.93% |
| Demographic Bias | 2.86 (2.82, 2.89) | 2.71 (2.66, 2.76) | <0.01 | 5.54% |
| Without Treatment Bias | 2.89 (2.86, 2.92) | 2.87 (2.70, 2.80) | <0.01 | 0.70% |
| Updated Information | 2.89 (2.87, 2.92) | 2.77 (2.72, 2.81) | <0.01 | 4.33% |
| Low to no Patient Risk | 2.90 (2.87, 2.93) | 2.78 (2.74, 2.82) | <0.01 | 4.19% |

Legend: non-normal distribution for all criteria according to the Shapiro–Wilk test.
Mann–Whitney (M-W) U was used for group comparisons.
The Percentage Relative Difference was calculated as ((Group A-Group B)/Group B) \* 100, representing the improvement of Group A compared to Group B.

### Validity score by specialty and type of question

Group A achieved significantly higher validity scores than Group B across all specialties and question types (both Mann–Whitney’ U and Kruskal-Wallis’ test p<0.01). Among specialties, the most significant relative improvement was in gynecology (11.48%), while the smallest was in psychiatry (1.75%) (Post Hoc Pairwise comparisons – DSCF method’s p-value was 0.005). Regarding question types, research questions showed the greatest improvement (9.47%), whereas general knowledge questions showed the least (2.95%) (DSCF’s p-value < 0.001). Full results are presented in Supplementary Material 5 (specialties) and Supplementary Material 6 (question types).

### Efficiency by groups

Table 6 presents the efficiency results for Groups A and B. The time between cases was determined using the records from the Quizziz platform. The times between questions were calculated in Jamovi V2.6.45, where each observation had an automatically assigned weight. On average, Group A took almost four minutes less than Group B. Regarding the number of searches, Group B conducted more than twice as many searches as Group A. The average number of searches per case was 14 for Group A and 30 for Group B, confirming the previous statement.

**Table 6.**
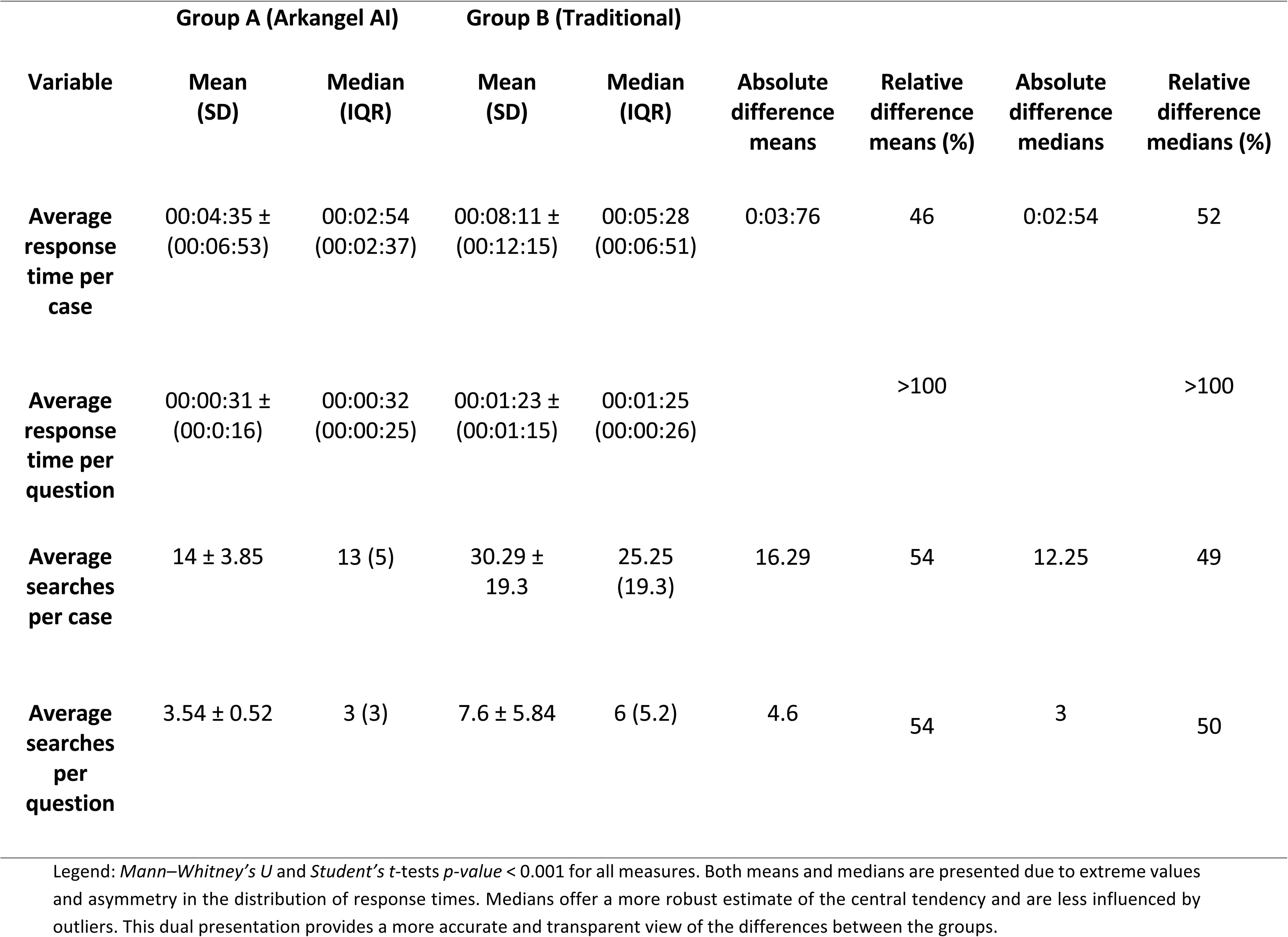
Efficiency results between groups: Average time and number of searches.

### Tool acceptability

Figure 2 shows the average scores obtained for the acceptability questions in Table 3, as ranked by subjects in group A. Perceived tool utility, likelihood of daily use, and recommendation to a colleague were scored 2.98, 2.88, and 2.92, respectively. The lowest score was observed for confidence in the model’s truthfulness, with 2.63. Total average tool acceptability is 2.85.

**Figure 2.**
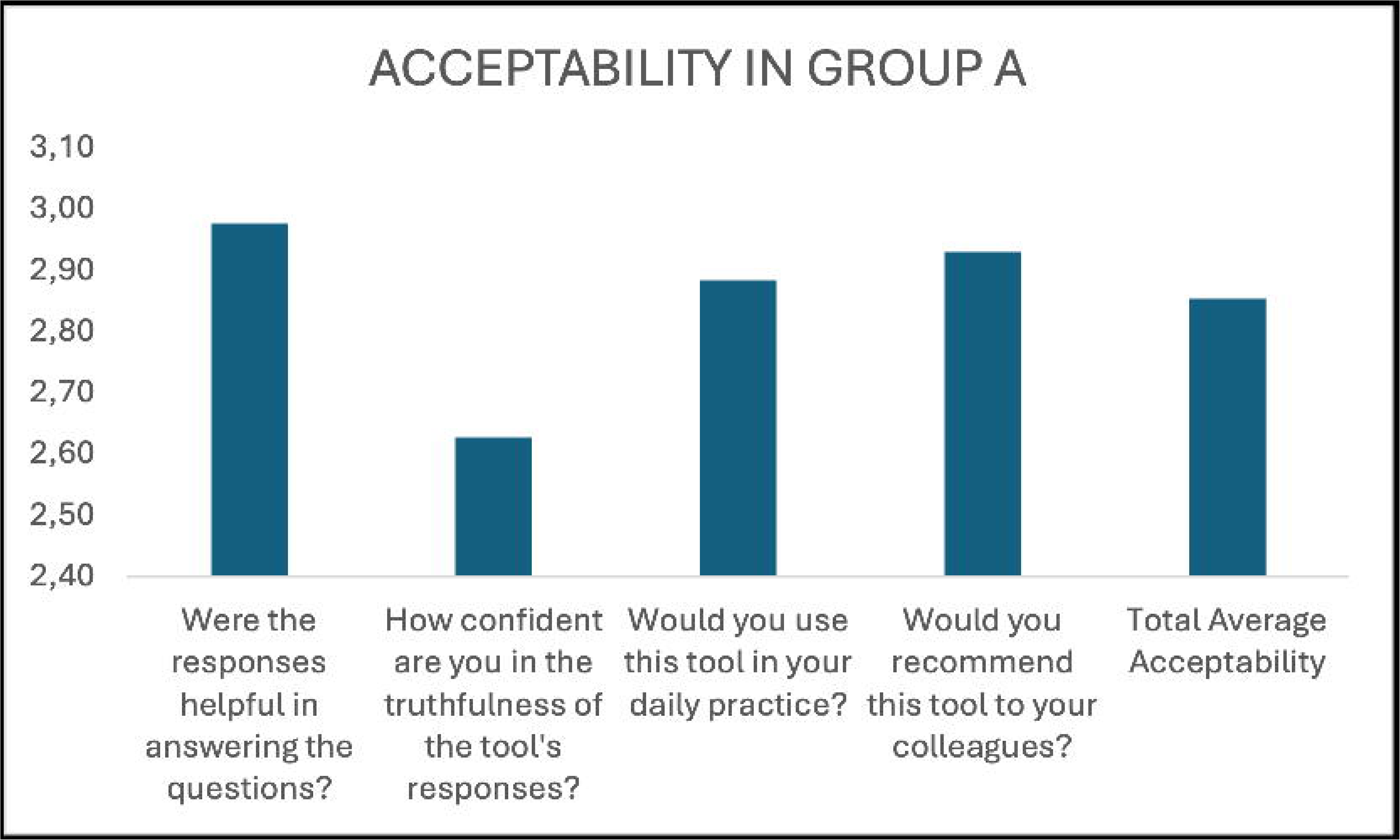
Previous and current system architecture.

**Figure 3.**
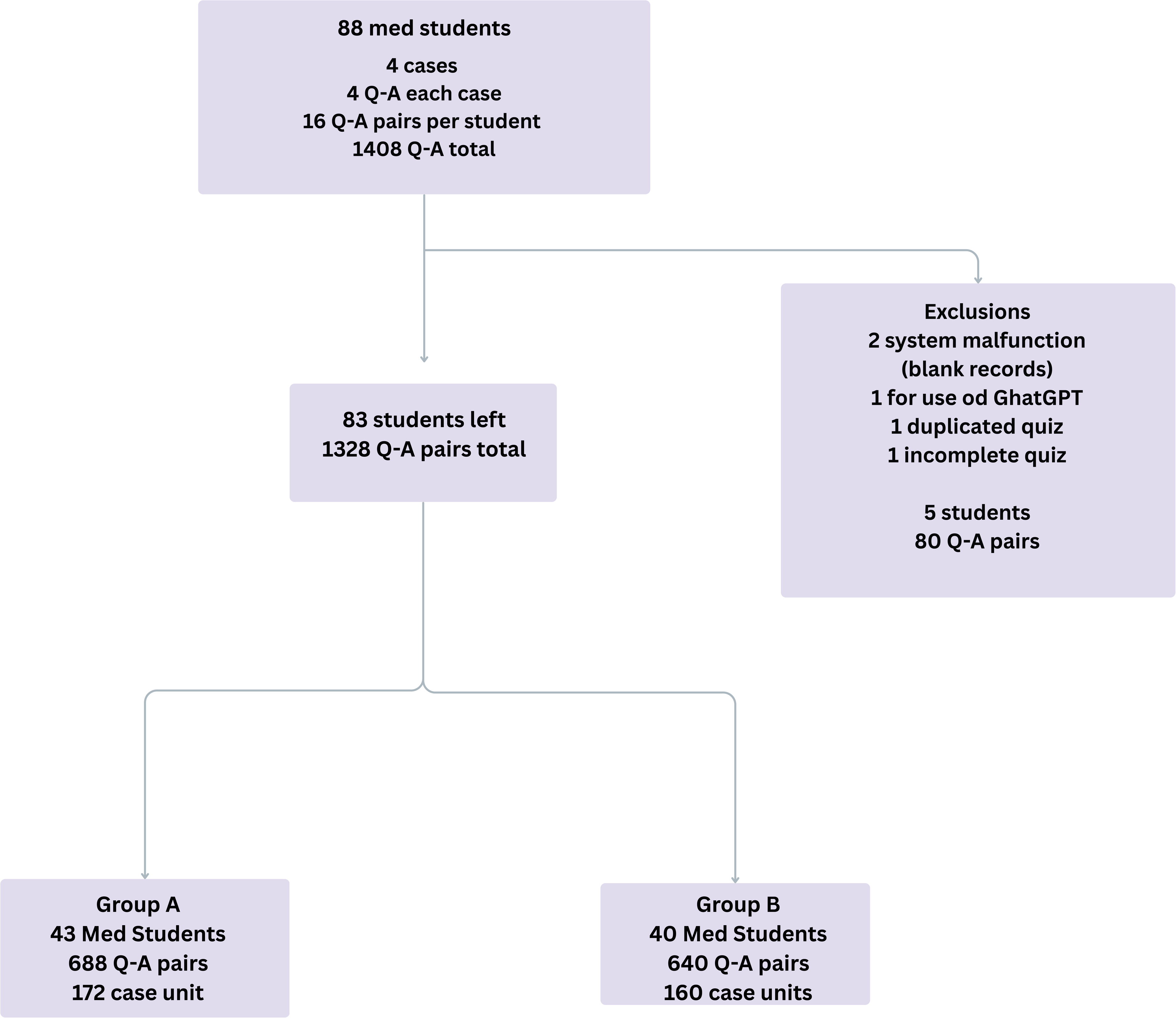
Average acceptability score for Arkangel AI (only for Group A)

## DISCUSSION

This is intended to be the first external validation of a series of studies assessing how ArkangelAI speeds up the process of finding clinically sound answers with the quality of a trained medical professional. Unlike traditional LLM evaluation approaches, which focus on standardized multiple-choice QA exams, this research takes a more analytical approach. It employs expert physicians’ knowledge as the standard of reference against which the human-in-the-loop agent’s answers are appraised regarding their quality and validity. It also compares the time taken using the different research methods.

The model’s performance can translate into abilities such as effective clinical information search, analysis, and synthesis skills, cognitive support, and evidence-based decisions. These abilities help the learning process and could act as pedagogical support, with AI digital medical education used as a tool to support evidence-based learning and early clinical reasoning (21–23).

Our findings support these claims and are consistent with recent studies showing how AI-based conversational assistants can improve the quality of clinical reasoning in medical students, provided they are integrated critically and supervised within training (21), and how they save time by obtaining faster responses, maintaining or augmenting their quality.

### Efficiency and Acceptability

The results demonstrated that students who used Arkangel AI solved clinical questions in significantly less time than the control group (median 2:54 vs. 5:28; relative reduction of 52%) and with fewer searches (3 vs. 6; relative decrease of 50%). Overall, the AI group required half the time and fewer than half the searches that the traditional search group. These differences were statistically significant (Mann–Whitney U test, p < 0.01). These suggest that Arkangel AI can enhance cognitive efficiency and enhance information seeking, essential skills for self-directed learning and evidence-based medical practice (24,25).

Overall acceptability was high (2.85 on a scale of 1 to 3), highlighting its usefulness and participants’ willingness to recommend it to their peers. However, the dimension with the lowest score was confidence in the accuracy of the responses (2.63), a finding consistent with that reported by Sakamoto et al. (26), among others (27), who document skepticism towards artificial intelligence systems in clinical and educational contexts. Rather than a weakness, this result represents an opportunity to critically assess how the supervised use of AI can strengthen digital competence, encouraging source verification, responsible interpretation of evidence, and understanding of the limitations of language models.

To promote informed trust, strategies of explainability and transparency should be demanded from any clinical LLM, such as direct visualization of the sources consulted, indication of the level of evidence, explicit reference to clinical guidelines, and breakdown of the reasoning followed by the model. This type of transparency not only improves the perception of reliability but also enhances the pedagogical value of the tool by transforming each interaction into an active exercise in learning and critical reflection.

### Validity

The validity of the responses, evaluated by independent experts in six dimensions (accuracy, consistency with guidelines, absence of demographic or therapeutic bias, timeliness, and safety), was consistently higher in the ArkangelAI group. The most significant increase in performance was observed in accuracy (10.78%, Mann–Whitney U p-value <0.01), followed by unbiased and updated information. These results indicate that AI can improve student responses by facilitating access to curated and up-to-date scientific literature, thereby reinforcing their training in evidence-based medicine. Total Average Validity Score was 2.84 for group A and 2.69 for group B (Mann–Whitney U p-value <0.01)

Furthermore, these findings align with recent language model evaluation frameworks that prioritize dimensions such as accuracy, consistency, bias, safety, and clinical relevance (28–31), supporting the potential for supervised use of ArkangelAI to strengthen the quality, reliability, and educational value of responses produced in medical training settings.

The validity construct was also explored by specialty (Gynecology, Psychiatry, Orthopedics, and Pediatrics) and type of question (diagnostic, management, research, and general knowledge). Group A consistently scored higher than Group B. Absolute and relative differences were statistically significant, with no overlap in 95% confidence intervals. All hypothesis tests revealed p<0.05 (Supplementary Materials 5 and 6).

The most significant performance improvement was for gynecology (relative difference 11.4%) compared to psychiatry (2.7%, Mann–Whitney U p-value <0.01), which is consistent with analysis describing that specialties with more patient interaction must respond to constant changes and will thus be the least "impacted" by AI, while clinical and surgical components will be more influenced)(25). Research questions had the greatest improvement with 8.65% (Mann–Whitney U p-value <0.01), compared to general knowledge with 2.87% (Kruskal-Wallis DSFC post hoc correction’s p-value <0.001, Supplementary Materials 8 and 9)

Although large language models (LLMs) have been widely described as clinical support tools, few studies have evaluated their impact in medical training contexts. Goh et al. assessed ChatGPT-4 in diagnostic reasoning using a randomized design with 50 physicians. They found no significant differences between groups, likely due to their limited sample size and focus exclusively on diagnostic performance(32).

In contrast, our study covered multiple domains of knowledge (diagnosis, management, research, and general knowledge) and showed significant differences in all dimensions evaluated. The differential performances observed in areas such as gynecology, pediatrics, or psychiatry are consistent with previous research showing variability in the accuracy of LLMs depending on the medical specialty (33).

Human evaluation frameworks have also been proposed to prioritize the accuracy, consistency, bias, and safety of responses generated by LLMs, seeking to ensure the validity and reliability of information in educational and clinical settings (34). In Latin America, although the adoption of AI in medical education is in its infancy, its potential for undergraduate learning contexts is particularly relevant (35). In this regard, the present study provides evidence on the pedagogical value of ArkangelAI as a tool to support evidence-based learning in medical students.

### Strengths and limitations

This study is one of the first external validations of an AI-based tool (Arkangel AI) applied to undergraduate students in a medical education context. Its randomized, double-blind design, clinical cases developed by external specialists, and independent expert evaluation provide methodological rigor and bias control. In addition, applying a multidimensional validity construct that included accuracy, concordance with guidelines, bias, timeliness, and safety offers a more comprehensive approach than traditional studies based solely on precision or accuracy of responses.

One methodological limitation was that responses were evaluated by a single specialist in each clinical area. This could introduce bias as the standard of reference is subject to their clinical educated opinion, and could be influenced by many human factors, like the specialist’s own experience with medical AI. In addition, it prevents the estimation of inter-rater variability, as each specialist only evaluated the case of their expertise. Therefore, even if the reviewers met experience criteria and were independent of Arkangel AI, some individual variability in scoring, inherent in human evaluations, cannot be ruled out. Nevertheless, human judgment remains the reference standard for LLMs (36). We plan to include more than one evaluator per specialty to assess the consistency and reproducibility of scores in future validations among medical staff.

Furthermore, the clinical cases used were fictitious, field-limited, and constrained to outpatient and low-complexity scenarios. Consequently, the results can only be applied to similar contexts for medical students at the clinical level. The results also reflect academic reasoning rather than clinical decision-making.

Another aspect is the potential conflict of interest, as Arkangel AI employs all authors. Although this relationship was transparently declared and mitigated by external evaluators and blind analysis, the need for independent validation by universities or hospitals to ensure replicability and scientific credibility is underscored. To address this further, we provide the project’s datasets for the replication of the analyses and results. We also describe the methodology and results as transparently as possible.

Since we did not use or divulge any patient data, we did not seek the approval of an ethics committee for this study, nor did we register it as a clinical trial (it does not fulfill the criteria). The cases were fictional and were not based on any patient. Every participant signed a consent form before entering the experiment.

### Educational implications and future lines of research

The evidence suggests that ArkangelAI can support students in developing efficient search skills, scientific information analysis, and initial clinical judgment, provided that its use is accompanied by faculty guidance and ethical standards (37). If properly framed, the incorporation of AI-based tools into medical education can become a catalyst for reflective learning and critical thinking (25)

Future research should explore the curricular integration of AI and evaluate its impact on students’ cognitive, ethical, and attitudinal competencies, which were not the aim of this study. Similarly, it will be essential to conduct multicenter external validations, expand the sample size, and analyze the effect of the tool in actual clinical practice through studies focused on physicians and specialists that confirm the reproducibility of these findings and consolidate the scientific basis for the ethical and responsible integration of artificial intelligence into medical education and practice. Furthermore, comparison with other AI models will enable Arkangel AI to position itself within the international landscape of educational and clinical tools based on large language models (LLMs) (38–41).

## Conclusion

LLM-supported methods appear to have a positive influence on effective clinical search without sacrificing, and even augmenting, the quality of answers. This is applicable at the clinical medical student level and for non-critical clinical reasoning. Validations, including graduated physicians and specialists, are needed to further understand the effect of LLMs in education and clinical practice.

## Data Availability

All data generated or analyzed during this study are included in this published article and its supplementary materials.

## Declarations

### Declaration of generative AI and AI-assisted technologies in the manuscript preparation process

While preparing this work, the author(s) used Jenni AI (42) and Anara (43) to structure the introduction section. After using this tool/service, the author(s) reviewed and edited the content as needed and took full responsibility for the content of the published article. We also used DeepL Translator (44) to translate the clinical cases into English and Grammarly (45) to help with the writing and editing processes. All scientific insights, data analysis, and conclusions were the sole responsibility of the authors.

### Ethics approval and informed consent

This study involved medical professionals who evaluated fictitious clinical cases designed exclusively for research purposes. All participants provided written informed consent prior to participation. No real patient data, medical records, or identifiable personal health information were used or disclosed at any stage of the study.

According to Resolution 8430 of 1993 of the Ministry of Health and Social Protection of Colombia (Ministerio de Salud y Protección Social de Colombia), which establishes the ethical standards for health research in Colombia, this study was classified as minimal risk research. Under Article 11 of this resolution, studies involving exclusively fictitious cases and no use of real patient data are exempt from mandatory review by an institutional ethics committee. Therefore, formal ethical approval by an Institutional Review Board was waived.

The study was conducted in accordance with the ethical principles outlined in the Declaration of Helsinki (2022) ^28^.

## Authorship Statement

All authors met the ICMJE criteria for authorship and approved the final manuscript.

## Acknowledgments

We would like to extend our sincere gratitude to the physicians who prepared the clinical cases and to those who participated in the evaluations, including Dr. Juliana Muñoz Restrepo, Dr. Laura Ovadia Cardona, Dr. Estefanía Bahamonde, Dr. Jorge Luis Molina Valderrama, Dr. Nataly Ávila, and Dr. Andrés David Álvarez. Finally, we appreciate the hundreds of physicians and medical students who responded to our invitation and completed their task with distinction, making this project possible.

## Data sharing

The deidentified datasets generated and analyzed during this study, including the questions, answers, and evaluation scores with their data dictionary, are provided in the supplementary material attached to this manuscript. The corresponding author can provide further details upon reasonable request.

## Contributions

NCV: Conceptualization, Methodology, Data curation, Formal analysis, Project administration, Supervision, Writing – review & editing

MCV: Data curation, Software Writing – review & editing IL: Data curation, Formal analysis, Investigation

JZ: Resources, Supervision, Writing – review & editing

LV: Resources, Writing – review & editing

