## Supplementary Material for "Clinical Med students’ validation of Arkangel AI: Are their responses any better when supported by the AI?"

**Supplementary Material 1.** Informed consent form signed by study participants (original version in Spanish), English translation obtained with ChatGPT.

**I**NFORMED CONSENT FOR THE STUDY:

Project Title:
 Validation study of a conversational agent
 (MedSearch, first version called Vitruvius) for real-time medical question answering
 VITRUVIUS: A conversational agent for real-time evidence-based medical question answering
 Principal Investigator: Natalia Castaño Villegas

I, ______________________, identified with ID __________________________,
 declare that:
 • I have understood the written and/or verbal information provided to me.
 • I have been able and am able to ask any questions I deem necessary about the study.
 • My participation is voluntary.
 • I allow the recording of my screen during the meeting, for the purpose of evidence registration for the research (if applicable).
 • I have been informed about:

1. The objectives of the study and its procedures.
2. The benefits and inconveniences of the process.
3. The procedure and purpose for which my personal data will be used and the guarantees of compliance with current legal regulations.
4. That I can revoke my consent at any time (without needing to explain the reason) and request the deletion of my personal data.
5. That I have the right to access and rectify my personal data.

• I authorize Arkangel AI or its designees to use the information I provide for analytical purposes, to prepare studies and publish them.
 • Arkangel AI is not violating any confidentiality agreements, personal copyrights, or intellectual property rights by using the information provided, regardless of whether the studies result in any publication or commercial product.

I CONSENT TO PARTICIPATE IN THIS STUDY

(Write YES or NO)

To confirm all of the above, I sign below:

Date: _____________________
 Signature: _____________________

Investigator's Name: Natalia Castaño Villegas
 Investigator's Signature:

SECTION FOR REVOCATION OF CONSENT

I,
 ……………………………………………………………………………………………
 Revoke my consent to participate in the process signed above.

Signature and Date of Revocation

**Supplementary Material 2.** Clinical cases and derived questions (translated from the original language, Spanish)

| **Specialty** | **Clinical case** |
| --- | --- |
| **Gynecology** | A woman in her early thirties, married and nulliparous, currently enrolled in a technical program and employed in a service-related job, seeks urgent evaluation for a one-month history of low mood, initial insomnia, loss of energy, and frequent nightmares (“I have nightmares almost every night”). She denies loss of appetite. She reports distractibility and long-standing passive thoughts of death beginning in adolescence. She had past episodes of suicidal ideation, the last one several years ago. She acknowledges emotional instability. In the past, she responded to frustration with self-injurious and aggressive behaviors (“I burned myself with cigarettes, punched walls, and tried to divert attention to something physical”). She also reports decreased libido.  Over the last two years, she has increased her alcohol consumption with loss of control (“we could drink for 2–3 days straight”), leading to functional impairment (“I’ve put myself at risk because I have blackouts”) and relationship difficulties. She also uses marijuana and tobacco regularly. She has been undergoing marital separation in the past month. She reports previous episodes with similar characteristics. She describes alternating affective episodes of increased energy and productivity followed by periods of slowing and low motivation. For the past two months she has also felt restless and anxious. She denies current or past hallucinations, delusional ideas, or persecutory thoughts.  Past history: Respiratory allergies. No surgeries. No known drug allergies.  Substance use: Cigarette use since adolescence, currently daily; vaping in recent years with limited benefit. Cannabis since late adolescence, now several times per week. Remote experimental use of cocaine, hallucinogens, and other substances; no recent use.  Family history: Relatives with depression, suicide, and substance use disorders. |
| **Question type** | |
| **Diagnosis** | What is the most likely diagnosis in this patient? |
| **Management** | What is the appropriate management for this patient according to current guidelines from the National Institute of Health (Instituto Nacional de Salud, INS)? |
| **Research** | What is the efficacy of HPV vaccination in preventing cervical cancer among women who did not receive the vaccine during adolescence? |
| **General knowledge** | What role does a positive family history play in this condition? |
| **Orthopedics** | Personal data: Female, adult in her early thirties, single, with incomplete formal education, currently a homemaker.  Chief complaint: “Persistent neck pain.”  History of present illness: The patient reports a history of cervical pain of approximately two months, with mechanical features, particularly noticeable during neck rotation and extension. The pain is intermittent and associated with a sensation of head instability and occasional dizziness, described as feeling as if her head were “floating” or “detached” from her body. Symptoms worsen with routine activities such as reading or watching television and are also exacerbated by rapid side-to-side neck movements. Recently, she has experienced pain radiating to the shoulders and occasional numbness in the fingers. Her medical background includes autoimmune disease and chronic musculoskeletal disorders that could contribute to these symptoms. The neck pain has increasingly limited daily activities, and usual analgesics have become less effective.  Past medical history: Autoimmune disease with inflammatory arthritis of large and small joints; irregular treatment with corticosteroids and disease-modifying agents. Metabolic disorder requiring insulin therapy, with irregular adherence. Chronic anemia with occasional transfusions. Chronic arthritis in wrists, knees, elbows, and ankles; mechanical hip pain with restricted mobility.  Physical examination: The patient is in acceptable general condition, alert and oriented. Musculoskeletal exam: Limited cervical spine rotation; swelling and warmth in some joints (elbows, metacarpophalangeal joints); muscle atrophy of the interossei in both hands. Spurling test: Negative, without clear radiculopathy. Cervical palpation: Localized tenderness with reduced range of head motion. Neurologic exam: No major motor or sensory deficits, though the patient describes mild paresthesia in the fingers.  Diagnostic studies: Cervical radiograph: Evidence of mild atlantoaxial (C1–C2) instability, more noticeable during movement. Cervical MRI: Ligamentous and degenerative changes in the atlantoaxial complex, suggesting instability. Electromyography: No evidence of significant cervical radiculopathy. |
| **Question type** | |
| **Diagnosis** | What is the most likely diagnosis from an orthopedic standpoint? |
| **Management** | What are the surgical indications in patients with atlantoaxial instability of the cervical spine? |
| **Research** | What is the efficacy and safety of cervical collars in patients with cervical spine pathology? |
| **General knowledge** | What recommendations and restrictions should be followed for therapeutic exercise and physical therapy in a patient with cervical spine pathology? |
| **Pediatrics** | A female infant of approximately two months of age, exclusively breastfed on demand, has a one-month history of frequent post-feed emesis and persistent straining. During the past week, the stools have shown streaks of blood. She has not received formula or complementary foods.  The perinatal history corresponds to a full-term newborn delivered by cesarean section without complications, with unremarkable prenatal care and normal obstetric ultrasounds. Birth weight was appropriate for gestational age, and the infant has received the immunizations recommended for her age.  Growth assessment shows a weight-for-length of −2 standard deviations, which represents a decline from a previous measurement of −1 SD. Weight-for-age is −1 SD and length-for-age is −0.5 SD, indicating faltering growth with downward crossing of curves, mainly affecting weight.  On physical examination, vital signs are within normal limits. The infant is well hydrated, alert, and interactive. Mucous membranes are moist, without jaundice, and the head is normocephalic. Chest expansion is symmetric, without retractions, and breath sounds are clear bilaterally. Cardiac auscultation reveals a regular rhythm with no murmurs. The abdomen is soft and non-distended, without palpable masses or organomegaly, and the umbilicus is normal. Palpation does not produce apparent pain. Perianal inspection shows a normal anus, without fissures or skin tags. The skin reveals mild eczema on the malar region and faint eczema in the antecubital fossae. Neurologic evaluation demonstrates normal tone and reflexes for age.  In summary, this is an exclusively breastfed infant with growth faltering, a one-month history of post-feed emesis and straining, and new-onset hematochezia, accompanied by mild eczema and otherwise unremarkable systemic findings. |
| **Question type** | |
| **Diagnosis** | What is the most likely diagnosis in this patient? |
| **Management** | What feeding options are recommended for the management of infants with this diagnosis? |
| **Research** | What is the effectiveness of maternal cow’s-milk elimination diets during exclusive breastfeeding for managing cow’s-milk protein allergy in infants? |
| **General knowledge** | How can proteins from the maternal diet trigger an allergic response in exclusively breastfed infants? |
| **Psychiatry** | A woman in her early thirties, married and nulliparous, currently enrolled in a technical program and employed in a service-related job, seeks urgent evaluation for a one-month history of low mood, initial insomnia, loss of energy, and frequent nightmares, which she describes as occurring almost every night. She denies loss of appetite. She reports distractibility and longstanding passive thoughts of death beginning in adolescence. She acknowledges a history of suicidal ideation, the most recent episode occurring several years ago. She describes herself as emotionally unstable and notes that in the past she reacted to frustration with aggression and self-injurious behavior, such as burning herself with cigarettes or punching walls. She also reports decreased libido.  Over the past two years she has increased her alcohol consumption with loss of control, sometimes drinking for two to three days consecutively. This pattern has resulted in functional impairment, including blackouts that she perceives as putting her at risk, and has contributed to marital conflict. She also reports regular use of marijuana and tobacco. She is currently undergoing marital separation and describes similar episodes in the past. She reports alternating periods of heightened affective activation, with a sense of being “revved up” and highly productive, followed by episodes of psychomotor slowing and low energy. For the past two months she has also experienced restlessness and anxiety. She denies current or past hallucinations, delusional ideas, grandiosity, or persecutory thoughts.  Her past medical history is notable for asthma and allergic rhinitis. She has no history of surgeries and no known drug allergies. Regarding substance use, she began smoking cigarettes in adolescence, with heavy use in the past five years; currently she smokes about ten cigarettes per day and has also been vaping in an attempt to reduce consumption, without success. Cannabis use began in late adolescence, with a daily pattern established in early adulthood; she currently consumes several joints per week, with the longest abstinence lasting only ten days, a few months ago. She reports remote experimental use of cocaine, hallucinogens, and LSD, but denies any use of MDMA.  Her family history is significant for mood disorders and substance use: one relative died by suicide, another had depression, and her parents have histories of alcohol and cannabis abuse. |
| **Question type** | |
| **Diagnosis** | What are the most likely diagnoses in this patient? |
| **Management** | What pharmacologic and non-pharmacologic management is recommended for this patient? |
| **Research** | What is the effectiveness of integrated treatment programs for bipolar disorder and substance use disorder in reducing relapses and improving functioning in adults? |
| **General knowledge** | How does bipolar disorder differ from major depression and from schizophrenia? |

**Supplementary Material 3.** Details on the dynamic modifications in the current ArkangelAI compared to its first versions

**Model Description**

We described the development and internal validation of Arkangel AI in a previous paper [1]. Improvements and iterations are ongoing, defining a fluid process in which efficiency, confidence, security, and satisfaction are priorities. Since then, the model has had two important updates.

The first one was for workflow classification. Our previous system had five LLMs to classify the information into workflows according to the type of question (diagnostic, clinical management, research, common knowledge) and retrieve relevant information. It generated four answers from the same information sources to refine the final answer. Workflow classification had an accuracy of 95% compared to the human standard. It did not favor a particular workflow (Kruskal-Wallis p-value >0.05), but the heavy processing impacted efficiency, with an average response time of 2.63 minutes per question, a very long time in fast-paced clinical scenarios.

Our current model still has a workflow-like structure. The user chooses between six options: clinical, pharma, differential, clinical plan, writer, and pharmaceutical industry. Each workflow uses a different LLM defined by the Machine Learning (ML) engineers, depending on the information sources (see Table 3 in the manuscript). The sources used by each line of question were determined using the recommended websites, applications, and resources that resulted from expert advice, model feedback, user experience, and consensus within the research team (see Supplementary Material 5).

The second modification was for the retrieval. For our previous model, retrieval was made using Google and PubMed's Application Programming Interfaces (APIs). Accuracy and Response Relevance were high (91.26% and 97%, respectively). Response relevance is a measurement in the RAGAS automated framework for retrieval augmented generation (RAG) evaluation, described by Shahul et al. in 2023, quantifying how much the answer addresses the question. However, Context Relevance measures how much of the results are essential for the response, revealing that only 80% of retrieved contexts were relevant. To improve retrieval efficiency, we used the AI search engine Tavily. This system works like a traditional Search API; the difference is that it uses a proprietary AI to search, scrape, filter, rank, and extract information from online sources (<https://docs.tavily.com/documentation/about>).

These two modifications made it possible to use resources and time more efficiently and improve response time by simplifying the multiple steps and LLMs needed to produce a refined answer, changing from a five-LLM structure to one with only two LLMs (see Figure 1 in the manuscript), and providing a faster retrieval using the paid application. Supplementary Material 5 provides more details about the architecture and connectivity of the Tavily retriever and the Arkangel workflows for reproducibility and transparency, together with a list of all Arkangel AI’s sources. The specific prompts are available upon reasonable request.

**Supplementary Material 4.** Retrieval pipelines

Our system uses the Tavily AI search engine for real-time semantic, contextual, and keyword-based retrieval. Unlike traditional search engines, Tavily analyzes multiple sources simultaneously and returns only the context needed to answer the user’s query, reducing latency (according to their website)

Access is provided through an API, which returns results in JSON format containing query, title, URL, and extracted content, among other metadata. A simplified example is shown below:

{

"query": User's query,

"images": [],

"results": [

{

"title": "", "url": "", "content": "", "score": 0.00,

"raw_content": null,

"favicon": ""

}

],

"auto_parameters": {

"topic": "general", "search_depth": "basic"

},

"response_time": 0.00,

"request_id": "0000"

}

Tavily’s retrieval quality was benchmarked with OpenAI’s SimpleQA dataset (4,326 questions), where it achieved 93.3% accuracy, compared with 41.6% using GPT-4.1 without retrieval.

For MedSearch, we generate three targeted clinical queries (two in English, one in the user’s language). Each query is sent to Tavily, which is restricted to a curated list of 37 trusted medical repositories (e.g., PubMed, Cochrane Library, ClinicalTrials.gov, JAMA, BMJ, WHO, FDA, EMA, etc.). Tavily is configured to return ≥5 articles per query. Earlier prototypes used six queries; the current version uses three for efficiency.

The complete list of biomedical repositoriesArkangel AI uses is provided as follows:

| **Workflow** | **Sources/Prompt** |
| --- | --- |
| **Clinical** | "https://pubmed.ncbi.nlm.nih.gov/",  "https://scholar.google.com/",  "https://www.cochranelibrary.com/",  "https://clinicaltrials.gov/",  "https://www.embase.com/",  "https://www.sciencedirect.com/",  "https://jamanetwork.com/",  "https://www.bmj.com/",  "https://journals.plos.org/plosmedicine/",  "https://onlinelibrary.wiley.com/",  "https://www.nih.gov/",  "https://www.scopus.com/",  "https://www.uptodate.com/",  "https://www.webofscience.com/",  "https://eric.ed.gov/",  "https://www.who.int/hinari/en/",  "https://www.medrxiv.org/",  "https://www.biomedcentral.com/",  "https://www.researchgate.net/",  "https://www.scielo.org/",  "https://www.semanticscholar.org/",  "https://portal.fiocruz.br/",  "https://www.insp.mx/",  "https://www.ins.gov.co/",  "https://www.isciii.es/",  "https://www.canada.ca/en/services/health.html",  "https://www.nhmrc.gov.au/",  "https://www.argentina.gob.ar/salud/",  "https://www.gov.uk/government/organisations/department-of-health-and-social-care",  "https://www.aihw.gov.au/",  "<https://www.santepubliquefrance.fr/>"  "<https://ashpublications.org/>"*,  "https://ascopost.com/"*. |
| **Pharma** | "https://dailymed.nlm.nih.gov/",  "https://www.drugs.com/"; |
| **Differential** | Performs a search based on the following prompt objective: "As a medical expert, generate a comprehensive clinical guideline and publications search given the patient's symptoms and medical history to determine that will support the differential diagnosis" |
| **Clinical Plan** | Performs a search based on the following prompt objective: "As a medical professional, conduct a comprehensive search supporting clinical guidelines and publications that will support a clinical plan for the given query" |
| **Writer** | Does not search. Designed for requests that only require writing support. |
| **Pharmaceutical Industry** | https://www.statista.com  https://www.researchandmarkets.com  https://www.marketwatch.com  https://www.fda.gov/  https://www.ema.europa.eu/  https://www.picscheme.org/ |

****Sources are updated frequently based on real-time feedback from users. The sources above correspond to those included at the time of the study.***

**Supplementary Material 5.** Total Average Validity Scores by Specialty

| **Specialty** | **Average Score** | | **Mann-Whitney's U** | **Relative difference (%)** |
| --- | --- | --- | --- | --- |
|  | **Group A** | **Group B** |  |  |
| **Pediatrics** | 2.87  (2.81, 2.92) | 2.67  (2.58, 2.75) | <0.01 | 7.49 |
| **Orthopedics** | 2.86  (2.82, 2.90) | 2.79  (2.74, 2.84) | <0.01 | 2.51 |
| **Psychiatry** | 2.91  (2.89, 2.93) | 2.86  (2.83, 2.88) | <0.01 | 1.75 |
| **Gynecology** | 2.72  (2.69, 2.79) | 2.44  (2.34 2.54) | <0.01 | 11.48 |

Legend: non-normal distribution for all criteria according to the Shapiro–Wilk test.
Mann–Whitney (M-W) U was used for group comparisons.
The Percentage Relative Difference was calculated as ((Group A-Group B)/Group B) * 100, representing the improvement of Group A compared to Group B.

**Supplementary Material 6.** Total Average Validity Scores by question type

| **Question Type** | **Average Score** | | **Mann-Whitney's U** | **Relative difference (%)** |
| --- | --- | --- | --- | --- |
|  | **Group A** | **Group B** |  |  |
| **Diagnostic** | 2.90  (2.86, 2.94) | 2.76  (2.70, 2.83) | <0.01 | 5.07 |
| **Management** | 2.78  (2.72, 2.85) | 2.64  (2.56, 2.73) | <0.01 | 5.30 |
| **Research** | 2.89  (2.84, 2.93) | 2.64  (2.56, 2.73) | <0.01 | 9.47 |
| **General Knowledge** | 2.79  (2.74, 2.84) | 2.71  (2.65, 2.77) | <0.01 | 2.95 |

Legend: non-normal distribution for all criteria according to the Shapiro–Wilk test.
Mann–Whitney (M-W) U was used for group comparisons.
The Percentage Relative Difference was calculated as ((Group A-Group B)/Group B) * 100, representing the improvement of Group A compared to Group B.

**Supplementary Material 8.** Post hoc Pairwise comparisons – DSCF method for the Kruskal-Wallis* test results by type of Question

| **Type of question 1** | **Type of question 2** | **W** | **p-value** |
| --- | --- | --- | --- |
| Diagnostic | General_Knowledge | -7.81 | < .001 |
| Diagnostic | Management | -5.50 | < .001 |
| Diagnostic | Research | -1.93 | 0.522 |
| General_Knowledge | Management | 2.26 | 0.379 |
| General_Knowledge | Research | 5.44 |  |
| Management | Research | 3.34 | 0.084 |

* Kruskal-Wallis p-value < 0.01

**Supplementary material 9.** Post hoc Pairwise comparisons – DSCF method for the Kruskal-Wallis* test results by Specialty

| **Type of question 1** | **Type of question 2** | **W** | **p-value** |
| --- | --- | --- | --- |
| ginecologia | ortopedia | 5.689 | < .001 |
| ginecologia | pediatria | 6.845 | < .001 |
| ginecologia | psiquiatria | 4.712 | 0.005 |
| ortopedia | pediatria | 2.582 | 0.261 |
| ortopedia | psiquiatria | 0.235 | 0.998 |
| pediatria | psiquiatria | -4.477 | 0.008 |

* Kruskal-Wallis p-value < 0.01
